# Cochrane Evaluation of (Semi-) Automated Review Methods (CESAR): Protocol for an adaptive platform study within reviews

**DOI:** 10.64898/2026.04.13.26350802

**Authors:** Gerald Gartlehner, Susan Banda, Max Callaghan, Jo-Ana Chase, Andreea Dobrescu, Angelika Eisele-Metzger, Ella Flemyng, Sean Gardner, Ursula Griebler, Bartosz Helfer, Pawel Jemiolo, Biljana Macura, Jan C. Minx, Anna Noel-Storr, Noosheen Rajabzadeh Tahmasebi, Amin Sharifan, Joerg J. Meerpohl, James Thomas

**Author notes:** These authors contributed equally to this work and share last authorship (Joerg J. Meerpohl, James Thomas). **Corresponding author:** Gerald Gartlehner; Dr. Karl Dorrek Strasse 30, 3500 Krems, Austria.

## Abstract

**Background:** Artificial intelligence (AI) has the potential to improve the efficiency of evidence synthesis and reduce human error. However, robust methods for evaluating rapidly evolving AI tools within the practical workflows of evidence synthesis remain underdeveloped. This protocol describes a study design for assessing the effectiveness, efficiency, and usability of AI tools in comparison to traditional human-only workflows in the context of Cochrane systematic reviews.

**Methods:** Members of the Cochrane Evaluation of (Semi-) Automated Review (CESAR) Methods Project developed an adaptive platform study-within-a-review (SWAR) design, modeled after clinical platform trials. This design employs a master protocol to concurrently evaluate multiple AI tools (interventions) against a standard human-only process (control) across three key review tasks: title and abstract screening, full-text screening, and data extraction. The adaptive framework allows for the addition or removal of AI tools based on interim performance analyses without necessitating a restart of the study. Performance will be assessed using metrics such as accuracy (sensitivity, specificity, precision), efficiency (time on task), response stability, impact of errors, and usability, in alignment with Responsible use of AI in evidence SynthEsis (RAISE) principles.

**Results:** The study will generate comparative data about the performance and usability of specific AI tools employed in a semi- or fully automated manner relative to standard human effort. The protocol provides a flexible framework for the assessment of AI tools in evidence synthesis, addressing the limitations of static, one-time evaluations.

**Discussion:** This study protocol presents a novel methodological approach to addressing the challenges of evaluating AI tools for evidence syntheses. By validating entire workflows rather than individual technologies, the findings will establish an evidence base for determining the viability of integrating AI into evidence-synthesis workflows. The adaptive design of this study is flexible and can be adopted by other investigators, ensuring that the evaluation framework remains relevant as new tools emerge.

## 1 Introduction

Systematic reviews and other forms of evidence synthesis are highly resource-intensive [1]. Because tasks such as title and abstract screening, risk-of-bias assessment, data extraction, and rating the certainty of evidence are susceptible to human error, they are typically performed manually by at least two independent reviewers. Although this practice enhances the correctness and rigor of review findings, it also substantially increases the time and cost required to produce and update evidence syntheses [2].

Artificial intelligence (AI) offers the potential to improve the efficiency of individual evidence-synthesis tasks and reduce the risk of human errors [3]. In this protocol, we define AI as “*advanced technologies that enable machines to do highly complex tasks effectively—which would require intelligence if a person were to perform them*,” following the definition provided in Responsible use of AI in evidence SynthEsis (RAISE) recommendations [4].

Recent advances in generative large language models (LLMs) have renewed interest in applying AI to evidence synthesis [5]. These models offer the potential to support more integrated and flexible (semi-) automated review workflows. Unlike earlier natural language processing (NLP) systems, LLMs can process long-form text without task-specific training and are adaptable to a wide range of review tasks. Notably, studies on literature screening [6–8] and data extraction [9–11] have reported promising results. However, the use of LLMs also introduces risks, including the generation of incorrect responses, fabrication of data or references, perpetuation of biases, and dissemination of misinformation [12]. Furthermore, achieving fully reproducible outputs may be more challenging with LLMs [13].

Many evidence-synthesis software providers and numerous start-ups have integrated AI to assist review teams with various tasks throughout the review process [14–16]. Comparative assessments within evidence-synthesis workflows are essential to determine which tasks AI tools perform worse than, similarly to, or better than human reviewers. Such evaluations help clarify which types of errors are introduced or mitigated when using AI tools for evidence synthesis. Methodologically, RAISE guidance outlines foundational principles for the transparent and scientifically sound evaluation of effectiveness of AI tools in evidence synthesis [4].

The rapid pace of AI-tool development for evidence synthesis necessitates new methodological approaches to assess effectiveness. In clinical research, platform trials gained significant momentum during the coronavirus disease 2019 (COVID-19) pandemic, driven by the urgent need to rapidly evaluate multiple potential therapies. Platform trials are adaptive clinical trial designs that use a single master protocol to assess multiple interventions simultaneously against a common control group within a specific disease area [17]. Unlike traditional trials, platform trials allow researchers to add or remove treatment arms based on interim results while the trial is ongoing, making them highly efficient and flexible [17].

Drawing on the principles of platform trials, we introduce an innovative methodological approach in this study protocol: an adaptive platform study within a review (SWAR), or adaptive platform SWAR. SWARs are embedded methodological studies conducted within systematic reviews to evaluate alternative methods or processes in real-world review settings [18]. By adapting the platform trial framework to the context of evidence synthesis, the adaptive platform SWAR enables prospective, flexible, and efficient evaluation of multiple AI tools or approaches within a single review process.

## 2 Objective

The **C**ochrane **E**valuation of (**S**emi-) **A**utomated **R**eview Methods (CESAR) project aims to compare the performance of conventional human-only approaches with semi-automated and fully automated methods for completing individual review tasks within ongoing Cochrane review updates. CESAR will evaluate the concordance between these approaches, quantify the human effort required, assess user experiences with AI tools, examine the stability of AI tool responses, and characterize both the types and impacts of errors associated with each process.

## 3 Research Questions

1. What is the comparative performance between conventional human-only methods and semi- or fully automated approaches for completing individual review tasks in Cochrane review updates?
2. How does the human effort required to complete individual review tasks compare conventional human-only and semi- or fully automated approaches?
3. What is the potential impact of errors from semi- or fully automated approaches on review findings and conclusions?
4. How consistent are AI tool outputs when repeatedly used for the same review tasks?
5. How do users perceive the usability and overall user experience of different AI tools when applied to specific review tasks within Cochrane review updates?

The following sections outline the core protocol elements (master protocol) applied to the adaptive platform SWAR. Task-specific methodological elements are detailed in the appendices (Appendix A: Title/Abstract Screening; Appendix B: Full-Text Screening; Appendix C: Data Extraction).

## 4 Master Protocol

CESAR evaluates (semi-)automated approaches for three core systematic review tasks: title and abstract screening, full-text screening, and data extraction. For each participating review, we will use a prospective, parallel-group design in which AI tools are applied for semi-automated (AI-assisted) and, where technically feasible, fully automated completion of these tasks. Conventional human-only processes on the same reviews, conducted in compliance with the Cochrane Handbook [19], will serve as the shared control group, providing a consistent baseline for comparison (see Figure 1).

**Figure 1.**
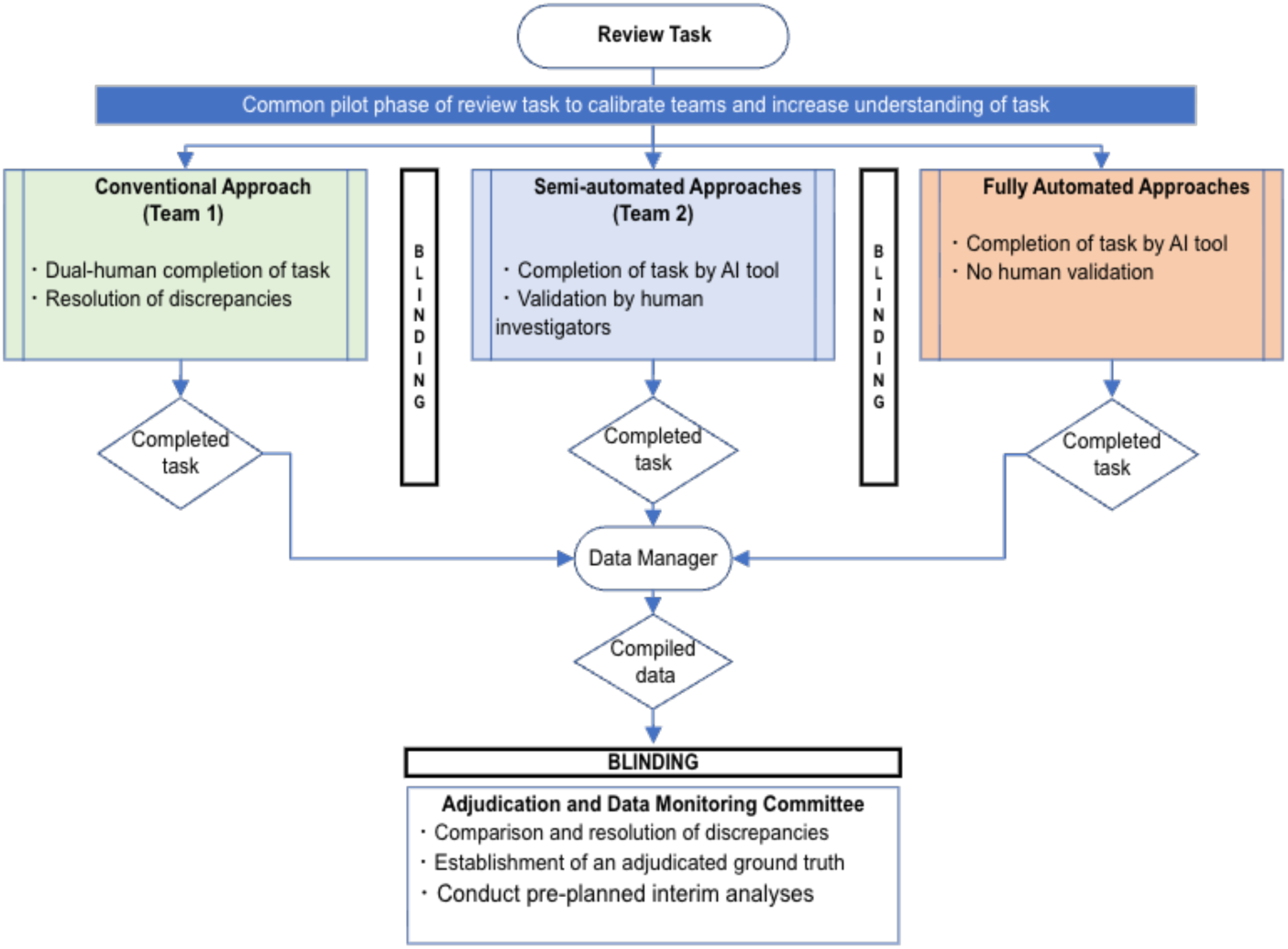
Study Flow Diagram Comparing Conventional, Semi-Automated, and Fully Automated Approaches to Systematic Review Tasks.

### 4.1 Selection of AI Tools

In November 2025, Cochrane invited developers of AI tools to submit proposals via a structured public form [20]. Proposals that met the initial screening for completeness were numerically scored on four criteria: adherence to the RAISE initiative [4]; transparency and interpretability of AI decisions; alignment with Cochrane’s mission and values; and compliance with data protection, copyright, and interoperability standards (e.g., RevMan) [21].

Shortlisted proposals were further evaluated by the Core Project Team and members of the joint AI Methods Group (https://methods.cochrane.org/ai/). Independent rankings from members of the joint AI Methods Group determined which two AI tools would initially enter the platform study, with others considered for inclusion if interim analysis led to discontinuation.

### 4.2 Selection of Reviews

We will select a convenience sample of fifteen ongoing updates of Cochrane intervention reviews of diverse topics based on the following criteria: 1) the review authors’ willingness to participate and the capacity to split into two teams with at least two members per team, 2) alignment of the review timeline with the study schedule, and 3) the expectation that the update will include at least one new study. Reviews requiring major methodological updates (e.g., rating the certainty of evidence) are not eligible. Each selected review will be integrated into CESAR after the corresponding review team has completed its literature searches. The review teams will establish and define the inclusion and exclusion criteria and then identify the data items for extraction.

### 4.3 Assembling Review Teams and Preparatory Work

For each review, we will establish two independent teams (hereafter referred to as Team 1 and Team 2) composed of members from the Cochrane review team conducting the update. While different individuals will comprise these teams, all members must possess review experience and a thorough understanding of the topic and review objectives. Team 1 (see Figure 1) will execute the conventional human-only approach, and Team 2 will implement two semi-automated approaches (one for each of the initially selected tools). If AI tools allow for fully automated approaches, members of the joint AI Methods Group (https://methods.cochrane.org/ai/) will handle automated tasks.

### 4.4 Common Pilot Phase

Before commencing the main evaluation, all participating teams will undergo a common pilot phase of the tasks for each review. This phase aims to align the teams, ensure consistent interpretation of instructions, and enhance familiarity with the tasks. During this phase, the teams conducting the semi-automated approaches will receive AI-tool–specific training. Insights gained from the pilot will be used to refine instructions and improve workflow comparability.

### 4.5 Review Task Completion

Teams assigned to different approaches will complete identical tasks under the same instructions but will be blinded to each other’s work to prevent performance bias. All completed tasks will be submitted to the data manager, who will compile a standardized, de-identified output matrix containing results from all study arms (conventional, semi-automated, and fully automated approaches). Group identifiers will be removed prior to submission to the Adjudication and Data Monitoring Committee (ADMC).

### 4.6 Reference Standards

Reference standards differ for each review task and are detailed in the respective appendices.

### 4.7 Data Adjudication and Interim Monitoring

The ADMC serves as an independent body that remains blinded for data adjudication. The committee composition will include a senior topical expert from each review team not involved in the specific tasks under evaluation alongside methodological experts. Within the adaptive framework, the ADMC has two primary mandates:

- Data adjudication
- Interim monitoring

#### 4.7.1 Data Adjudication

The ADMC will systematically assess and resolve discrepancies between conventional, semi-automated, and fully automated approaches using pre-specified adjudication criteria. For data extraction, the ADMC will establish the reference standard using a structured adjudication process:

- The ADMC will resolve discrepancies using a consensus-based method and original source documents.
- The ADMC will determine which approach made an error and assess its severity based on the potential impact on the review’s results and conclusions (see Table 1).
- All adjudication decisions, including dissenting opinions and their rationale, will be documented in a standardized adjudication log to ensure transparency and reproducibility.

**Table 1.**
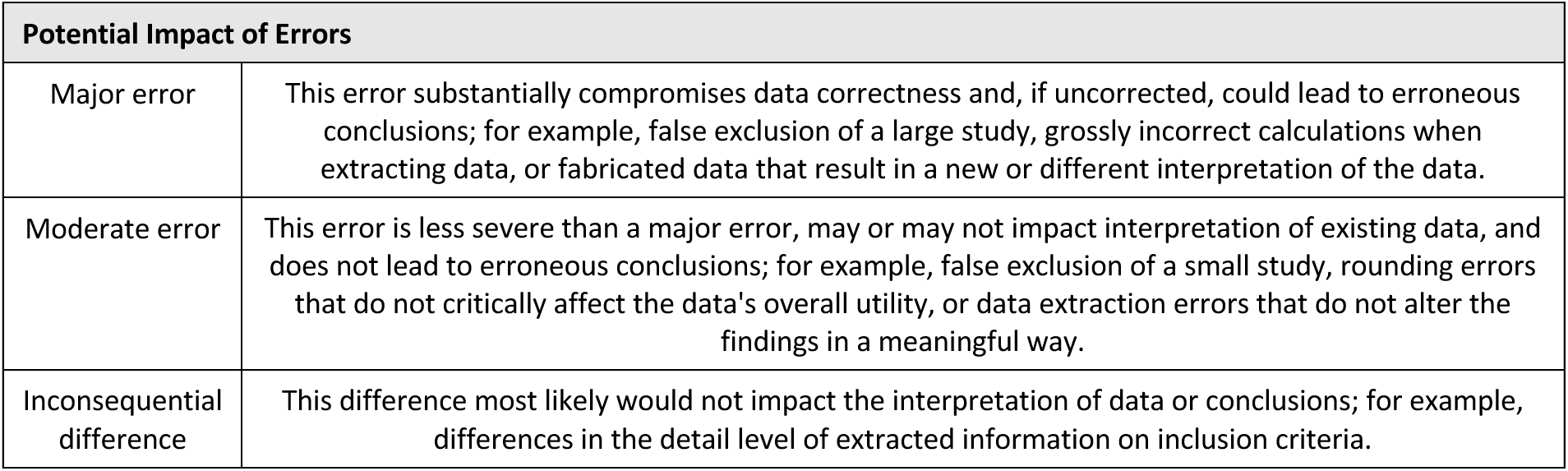
Classification of the potential impact of errors (adapted from Gartlehner et al. [22])

#### 4.7.2 Interim Monitoring

The ADMC will conduct at least one pre-planned interim analysis to evaluate whether any AI tool should be discontinued for futility (failing to meet minimum performance thresholds) or harm (introducing systematic errors that could compromise review validity). A formal interim analysis will be conducted for each AI tool after the completion of five reviews (33% information fraction). The ADMC will be unblinded for interim analysis.

Each AI-supported approach will be evaluated against performance thresholds using a hybrid decision rule that considers both point estimates and confidence intervals (Table 2). Specifically, an AI tool will be considered for discontinuation if:

- the observed point estimate falls below the futility boundary, indicating unambiguous underperformance; or
- the upper limit of the 95% confidence interval falls below a non-inferiority margin, indicating failure to meet acceptable performance even under optimistic assumptions.

**Table 2.**
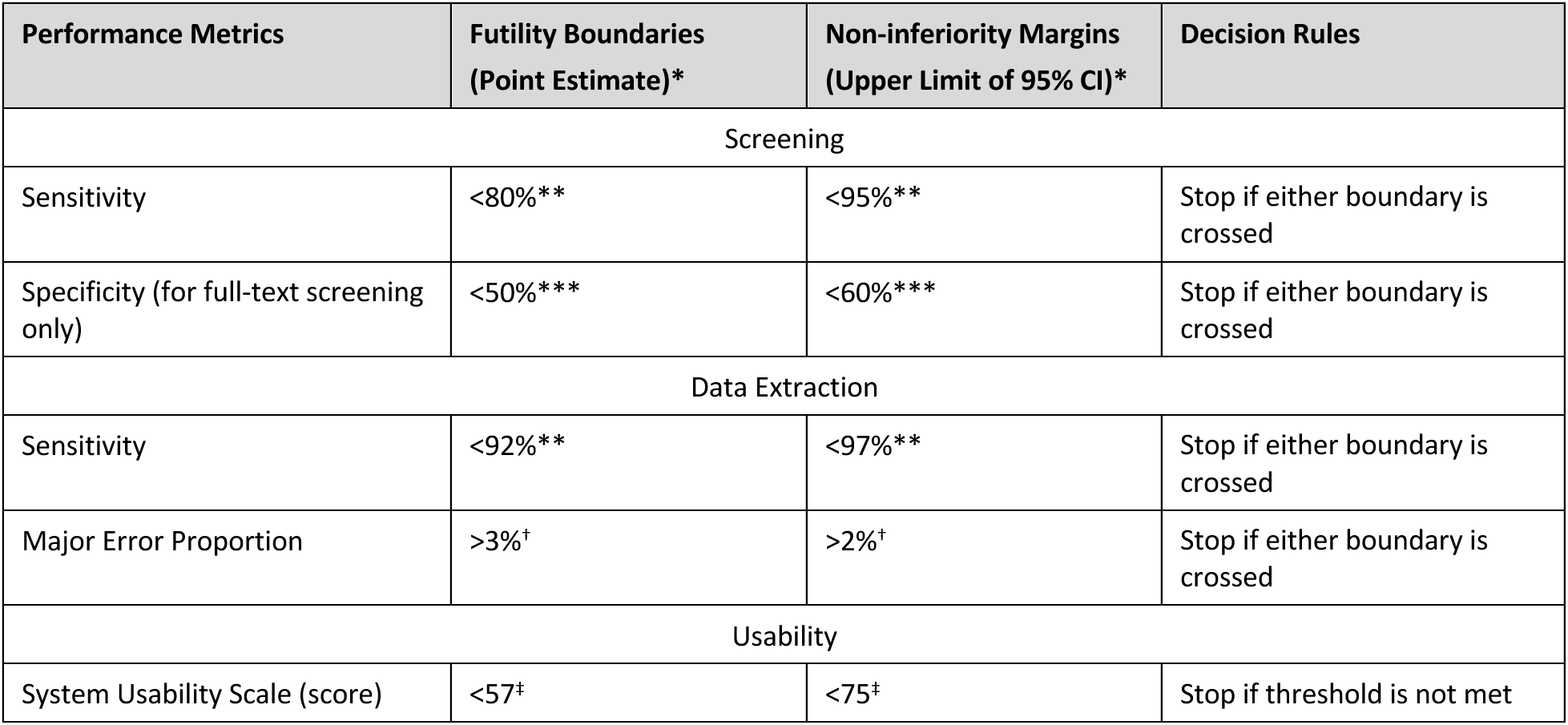
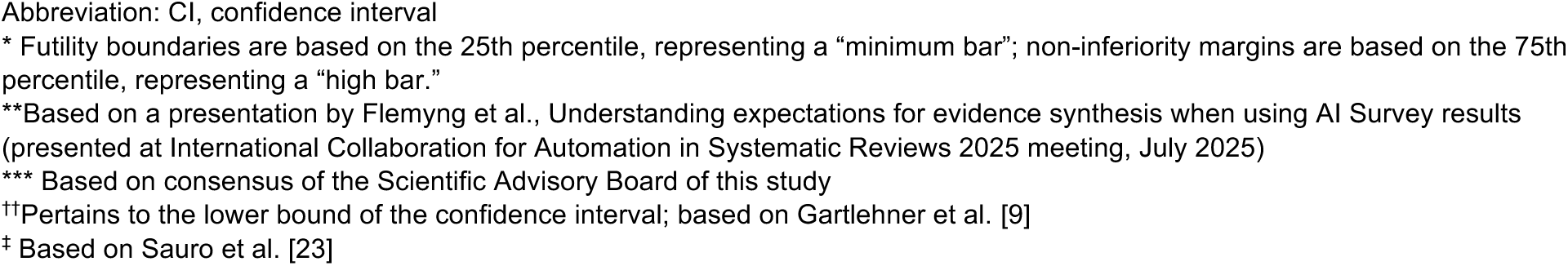
Stopping Boundaries and Decision Rules for Interim Analyses.

| Performance Metrics | Futility Boundaries<br>(Point Estimate)* | Non-inferiority Margins<br>(Upper Limit of 95% CI)* | Decision Rules |
| --- | --- | --- | --- |
| Screening |  |  |  |
| Sensitivity | <80%** | <95%** | Stop if either boundary is crossed |
| Specificity (for full-text screening only) | <50%*** | <60%*** | Stop if either boundary is crossed |
| Data Extraction |  |  |  |
| Sensitivity | <92%** | <97%** | Stop if either boundary is crossed |
| Major Error Proportion | >3%<sup>†</sup> | >2%<sup>†</sup> | Stop if either boundary is crossed |
| Usability |  |  |  |
| System Usability Scale (score) | <57<sup>‡</sup> | <75<sup>‡</sup> | Stop if threshold is not met |

Abbreviation: CI, confidence interval
\* Futility boundaries are based on the 25th percentile, representing a “minimum bar”; non-inferiority margins are based on the 75th percentile, representing a “high bar.”
\*\*Based on a presentation by Flemmyng et al., Understanding expectations for evidence synthesis when using AI Survey results (presented at International Collaboration for Automation in Systematic Reviews 2025 meeting, July 2025)
\*\*\* Based on consensus of the Scientific Advisory Board of this study
††Pertains to the lower bound of the confidence interval; based on Gartlehner et al. [9]
‡ Based on Sauro et al. [23]

#### 4.7.3 Decisions Regarding Changes in the Study Design

Decisions regarding modifications to the study design, including the selection of review tasks, the AI tools under evaluation, and the total number of included reviews, will be governed by the adaptive framework and overseen by a Cochrane Oversight Committee, the ADMC, and the principal investigator (GG). This structure allows the study to remain flexible, responding to interim findings and AI tool developments while maintaining methodological rigor. Specifically, the ADMC will review preliminary analyses to determine whether a tool’s performance warrants its removal from the platform study. Should an AI tool be discontinued, the next AI tool on the ranked shortlist will be enrolled in the study for subsequent reviews, ensuring a continuous and efficient evaluation process. Similarly, adjustments to the number of reviews or the scope of review tasks will accommodate emerging AI capabilities or enhance the precision of performance estimates. All modifications will be documented with a clear rationale and timestamps to maintain a transparent and auditable trail throughout the study.

### 4.8 Sample Size Considerations

Because this study is exploratory rather than confirmatory in nature, no hypotheses will be tested, and no formal sample size calculations will be performed. This approach allows for flexible, descriptive, and comparative analyses and provides the opportunity to explore multiple aspects of semi- and fully automated review processes. The fifteen review updates are a convenience sample, selected based on information from the Cochrane editorial team about reviews expected to be completed within the CESAR study timeframe.

### 4.9 Unit of Evaluation and Performance Metrics

We will use individual review tasks as the unit of evaluation (rather than entire reviews encompassing multiple review tasks) because this approach provides more detailed insight into the performance of AI tools. Assessing tasks individually allows the use of a consistent baseline for the conventional, the semi-automated, and the fully automated approaches for each new review task, thereby preventing errors that propagate from one task to the next within a review.

The choice of performance metrics will depend on the specific review task under evaluation and will be presented in more detail in the appendices on the individual tasks. In general, we will follow the Responsible use of AI in evidence SynthEsis-2 (RAISE 2) recommendations for performance metrics [24].

#### 4.9.1 Accuracy Metrics

The assessment of accuracy metrics will vary by review task and is described in more detail for each review task in the appendices.

#### 4.9.2 Stability Metrics

We will assess the stability of an AI tool’s output if an Application Programming Interface (API) is available. We will conduct ten independent iterations of fully automated tasks within a single day. For all tasks, our primary focus will be on sensitivity (recall). This approach will provide an initial estimate of variability in sensitivity arising from stochastic model behavior. During these iterations, no human verification will be performed, allowing assessment of the intrinsic stability of the fully automated approach. This process differs from our primary semi-automated approach, in which human reviewers will verify all AI screening decisions.

#### 4.9.3 Efficiency Metrics

We will assess time on task, defined as the total time required to complete a review task from initiation to verified output. For conventional approaches, this includes all steps typically performed by trained reviewers for calibration exercises, title/abstract and full-text screening, and data extraction. For semi-automated approaches, the same steps above are covered, plus time for: 1) tool configuration, 2) human verification of tool output and error correction, and 3) any necessary iteration or refinement. We will exclude initial training time for reviewers learning to use AI tools from the primary analysis but report it separately as implementation overhead. Time measurements will be recorded by reviewers using a time-tracking app. We will calculate the mean time on task with standard deviations.

For data extraction, a member of the Cochrane study team will measure the time required to convert the (semi-)automated output into a data format compatible with RevMan.

#### 4.9.4 Usability Metrics

Usability will be assessed using an electronic survey for each AI tool after completion of each review task. We will apply the System Usability Scale (SUS), a validated 10-item questionnaire that provides scores from 0 to 100 [25]. Scores below 70 indicate products needing scrutiny and improvement, whereas scores above 90 are considered superior [26]. The SUS has been successfully used to evaluate digital tools and systems [23]. Alongside usability, we will assess constructs relevant to the adoption of AI tools, including user satisfaction, trust in automation, and perceived risk. For trust in automation, we will use a subscale from the “Trust in Automation” questionnaire [27]. In addition, qualitative feedback will be collected through open-ended questions and analyzed thematically to identify common usability barriers and facilitators. Quantitative findings will be interpreted alongside qualitative feedback, as numerical scores alone may not capture AI-specific usability concerns. The survey will be conducted anonymously, and responses will not be traceable to individual review team members. An ethics waiver for the questionnaire was granted by the Ethics Committee of the University for Continuing Education Krems.

#### 4.9.5 Risk Assessment and Error Analysis

In accordance with Paper 2 of the RAISE guidance, we will assess the impact of errors, focusing on three outcomes: 1) error severity scoring (which will be performed by the ADMC, see Table 1), 2) proportions of missed critical information, and 3) changes in evidence-synthesis results [24].

The definitions of missed critical information differ by task and are detailed for each task in the appendices. Changes in evidence-synthesis results will be evaluated by recalculating effect estimates for the main outcomes in each review, either by excluding missed studies or by using erroneous data. This evaluation will address both the cumulative impact of all errors introduced by automating a task and the marginal impact of individual errors using leave-one-out sensitivity analysis [28].

For each pair of meta-analyses, we will assess the agreement between the recalculated and original effect estimates using Lin’s concordance correlation coefficient (CCC). We will first convert all dichotomous main outcomes to standardized mean difference (SMD) to have a common metric for all studies [29]. Lin’s CCC measures both co-variance and accuracy, with values close to +1 indicating stronger overall agreement and values near 0 or negative indicating poor agreement. We will also investigate the statistical significance of differences between recalculated and actual effect estimates using z-scores, which quantify the difference relative to the combined uncertainty of the two estimates.

Finally, we will use methods similar to Nussbaumer-Streit et al. [30] to assess whether differences in effect estimates imply a change in review conclusions. Specifically, a changed conclusion will be defined as cases where review authors report: 1) the same conclusion with less certainty, 2) a different conclusion (e.g., opposite direction), or 3) the inability to draw a conclusion.

### 4.10 Statistical Analysis

The statistical approach for this study will be primarily descriptive, reflecting its exploratory objectives. We will not conduct formal inferential statistics for direct comparisons between individual approaches.

Continuous outcomes, such as time on task, will be summarized using means, standard deviations, and 95% confidence intervals to characterize the distribution and uncertainty of measurements. For dichotomous outcomes and diagnostic performance metrics (e.g., sensitivity, specificity, and precision), we will report proportions alongside 95% Clopper–Pearson confidence intervals. This exploratory design is intended to generate robust descriptive insights that will inform future hypothesis-driven research and provide a foundation for methodological guidance on the responsible integration of AI tools within systematic review workflows.

### 4.11 Data Management

A data manager will oversee the data collection process. We will store data electronically in Excel datasheets in secure SharePoint folders managed by Cochrane. All investigators will have access to the data. The final prompts, model configuration settings, and human- and model-generated data outputs will be saved and shared upon request as part of dissemination. Likewise, we will make the final study data available upon request. Anyone wishing to withdraw their involvement in the project and delete their data will be able to do so. Anonymized and unidentifiable quotes or responses will only be used in project outputs, which may be public, with written permission.

### 4.12 Ethical Considerations

The study is primarily aimed at advancing knowledge and understanding in automated review task completion. For the usability survey, an ethics waiver was granted by the Ethics Committee of the University for Continuing Education Krems. For the Cochrane systematic review teams, corresponding authors will provide written consent on behalf of all authors to be part of the study and for Cochrane to use the data required for the study, including published and unpublished review data. The Cochrane review teams will use the AI tools, and any data inputs or outputs remain the responsibility and property of the Cochrane review teams, as is standard practice with AI-tool use by researchers. We will use AI tools that do not train on uploaded PDFs or user interactions, ensuring that the intellectual property rights of copyrighted study reports and review team data are fully respected.

## 5 Discussion

Validating AI tools within evidence-synthesis workflows is necessary to determine whether these technologies deliver practical value under real-world review conditions rather than only in controlled experimental settings. CESAR addresses this need by introducing an adaptive platform SWAR design that enables the concurrent, prospective evaluation of multiple AI tools across different evidence-synthesis tasks. By adapting the logic of platform trials from clinical research, this design offers an important methodological advance over static SWAR evaluations. In particular, it allows tools to be added, modified, or removed as technologies evolve while preserving a structured comparative framework. This flexibility is especially important in a field where AI systems change rapidly and results of conventional evaluation designs risk becoming outdated before they can be implemented.

A major strength of this study design is its orientation as a workflow validation study. Unlike model validation studies, which typically assess performance on pre-existing benchmark datasets under tightly controlled conditions, workflow validation embeds the AI tool directly into ongoing review processes [22]. This provides a more meaningful assessment of effectiveness, efficiency, usability, and error consequences in practice. The prospective nature of the design further strengthens the study by reducing the risk of data contamination, which may occur when evaluation datasets overlap with the data used to train LLMs [31]. Because developers rarely disclose the contents of training datasets, the use of ongoing unpublished reviews offers an important safeguard against inflated estimates of performance.

Another strength is the explicit incorporation of RAISE principles into the evaluation framework [4]. Accuracy alone is insufficient for judging whether an AI tool can be responsibly integrated into evidence synthesis. By also evaluating usability, response stability, time on task, and the impact of errors, the study recognizes that implementation decisions depend on a broader set of considerations than performance metrics alone.

The study adopted a rigorous approach to defining the reference standard. Due to variability in human inclusiveness when screening titles and abstracts, human decisions alone do not provide an optimal reference standard. By using the final set of included study reports as the reference standard, the study minimized the risk of variability across different reviews. Furthermore, instead of presuming that human-only data extraction is inherently reliable, any discrepancies between human-only and AI-assisted data extraction were adjudicated against the original study reports by blinded assessors. This process reduces benchmark bias resulting from imperfections in the human comparator and enables a more equitable assessment of all approaches.

Several limitations should be acknowledged. First, when using the final set of included study reports as the reference standard for title and abstract screening, it is not possible to reliably distinguish false positives from true negatives in the semi- and fully automated approaches. Consequently, metrics such as specificity, precision, and overall accuracy cannot be calculated. Due to time constraints, we limited the number of extracted data items to a maximum of 35, which does not represent the full range of data typically extracted in systematic reviews. Additionally, adjudicating discordant extractions and classifying error type and severity inevitably involve subjective judgment. Although employing two independent adjudicators and a third reviewer to resolve disagreements helps reduce arbitrariness, some inconsistency is unavoidable. Furthermore, by adjudicating only discordant extractions, concordant but jointly incorrect extractions may go undetected. While such cases may be rare, they remain a potential source of bias.

Overall, CESAR provides a rigorous and forward-looking framework for evaluating AI in evidence synthesis. Its chief contribution is methodological: it moves assessment beyond static benchmarks and toward prospective, adaptive, workflow-based validation that is better aligned with real-world implementation. This study design is readily adoptable by other teams seeking to validate AI tools within evidence-synthesis workflows.

## 6 Appendix A: Title/Abstract Screening

The evaluation of AI-assisted and fully automated title and abstract screening will adhere to the master protocol outlined above. Figure A1 illustrates the methodological framework for assessing this review task. The subsequent sections detail the methodological approaches that are specific to evaluating title and abstract screening.

**Figure A1.**
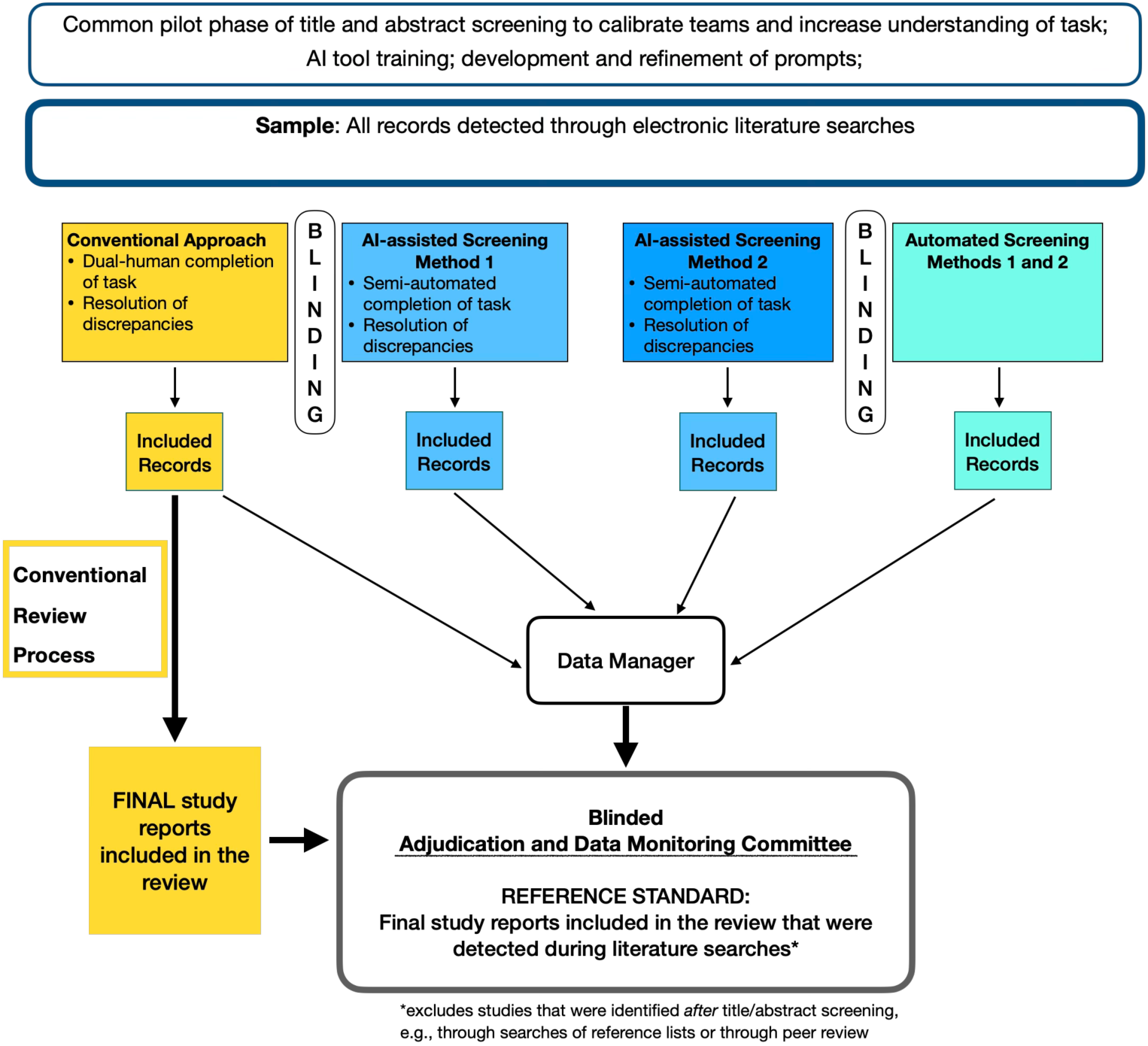
Methodological Framework for Title and Abstract Screening Performance.

### 6.1 Sample

The sample for title and abstract screening will be all records that were detected through electronic literature searches after deduplication.

### 6.2 Unit of Analysis

The unit of analysis is the record retrieved from electronic literature searches that is subject to title and abstract screening to determine whether it meets the inclusion criteria.

### 6.3 Reference Standard

The reference standard will consist of the *final included study reports identified through electronic literature searches* in the completed review. Study reports that meet the inclusion criteria but are discovered only at later stages (e.g., through reference list searches or during peer review) are excluded from this standard. This approach minimizes human variation during the title and abstract screening phases, where inconsistencies can be considerable. However, it introduces verification bias, as only abstracts that pass the initial human-led screening proceed to full-text review for confirmation. Consequently, this affects the accuracy metrics and the evaluation of false exclusions (false negatives) by human reviewers. Any false exclusions made by human reviewers that are not corrected in subsequent stages, such as reference list screening or peer review, remain undetected.

### 6.4 Performance Metrics

#### 6.4.1 Accuracy Metrics

Because of the chosen reference standard (final included study reports), the number of false positive decisions (records incorrectly marked for inclusion that were later excluded at full-text level) and the number of true negative decisions (records correctly excluded) of the semi- and fully automated approaches cannot be reliably distinguished. Consequently, performance metrics such as accuracy, specificity, and precision cannot be estimated. Sensitivity (recall), therefore, becomes the primary reliable metric, which is not affected by verification bias:

- *Sensitivity:* The proportion of the final included study reports that a title/abstract screening approach correctly identified

Exhibit A1 defines relevant accuracy metrics and components of the diagnostic 2 × 2 table for title and abstract screening. Proportions will be calculated with 95% Clopper–Pearson confidence intervals. For the fully automated screening approach, we will calculate the median sensitivity and the median proportion of included records across all iterations to assess response stability.

In addition, we will assess the following:

- *Proportion of included records*: The percentage of all screened records that are marked as “include” for full-text review

### Exhibit A1. Definitions of Accuracy Metrics for Title and Abstract Screening

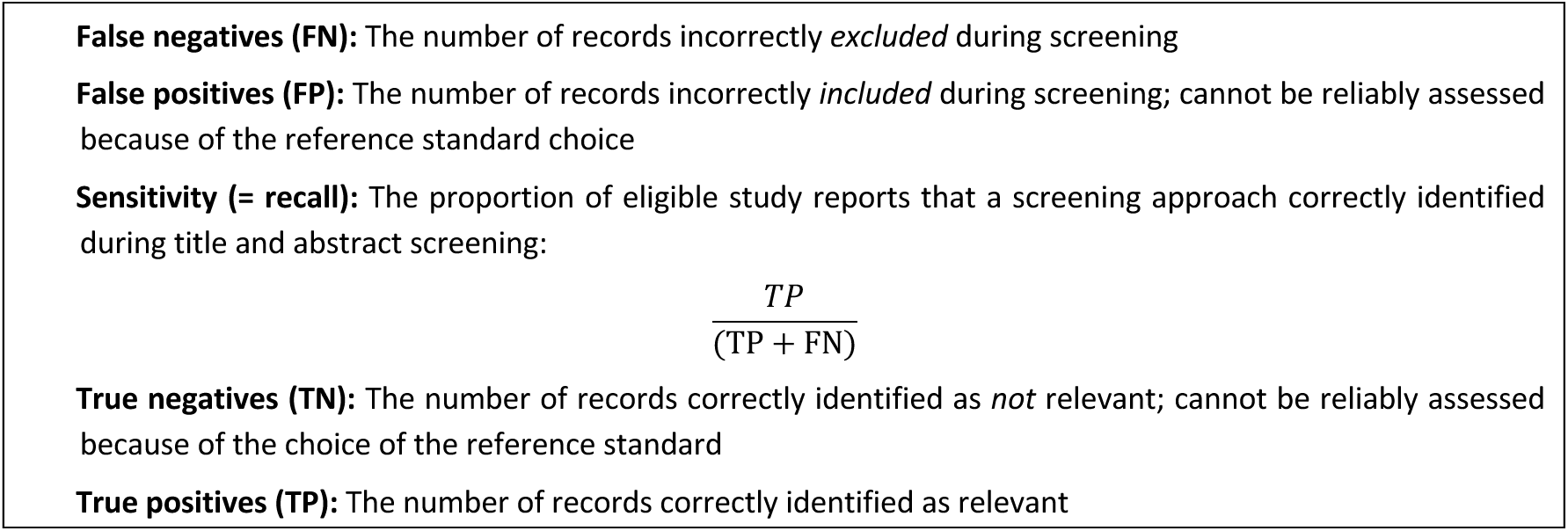

#### 6.4.2 Response Stability

Refer to the master protocol.

The assessment of response stability will focus on sensitivity (recall). For each run of the tool, true positives and false negatives will be identified at the individual review level. To assess reproducibility, we will compare the sets of false negatives across runs to determine whether the same study reports are consistently falsely excluded by the fully automated approach. The overlap between false negative sets will be quantified by calculating the mean pairwise Jaccard similarity [32].

#### 6.4.3 Efficiency Metrics

Refer to the master protocol.

#### 6.4.4 Usability Metrics

Refer to the master protocol.

#### 6.4.5 Risk Assessment and Error Analysis

Refer to the master protocol.

For title and abstract screening, missed critical information is defined as the proportion of eligible study reports incorrectly excluded during title and abstract screening. Changes in evidence-synthesis results will be analyzed by recalculating meta-analyses without incorrectly excluding study reports at this stage.

## 7 Appendix B: Full-text Screening

The evaluation of semi- and fully automated full-text screening will adhere to the master protocol outlined above. Figure B1 graphically illustrates the methodological framework for assessing this review task. The subsequent sections detail the methodological approaches that are specific to evaluating full-text screening.

**Figure B1.**
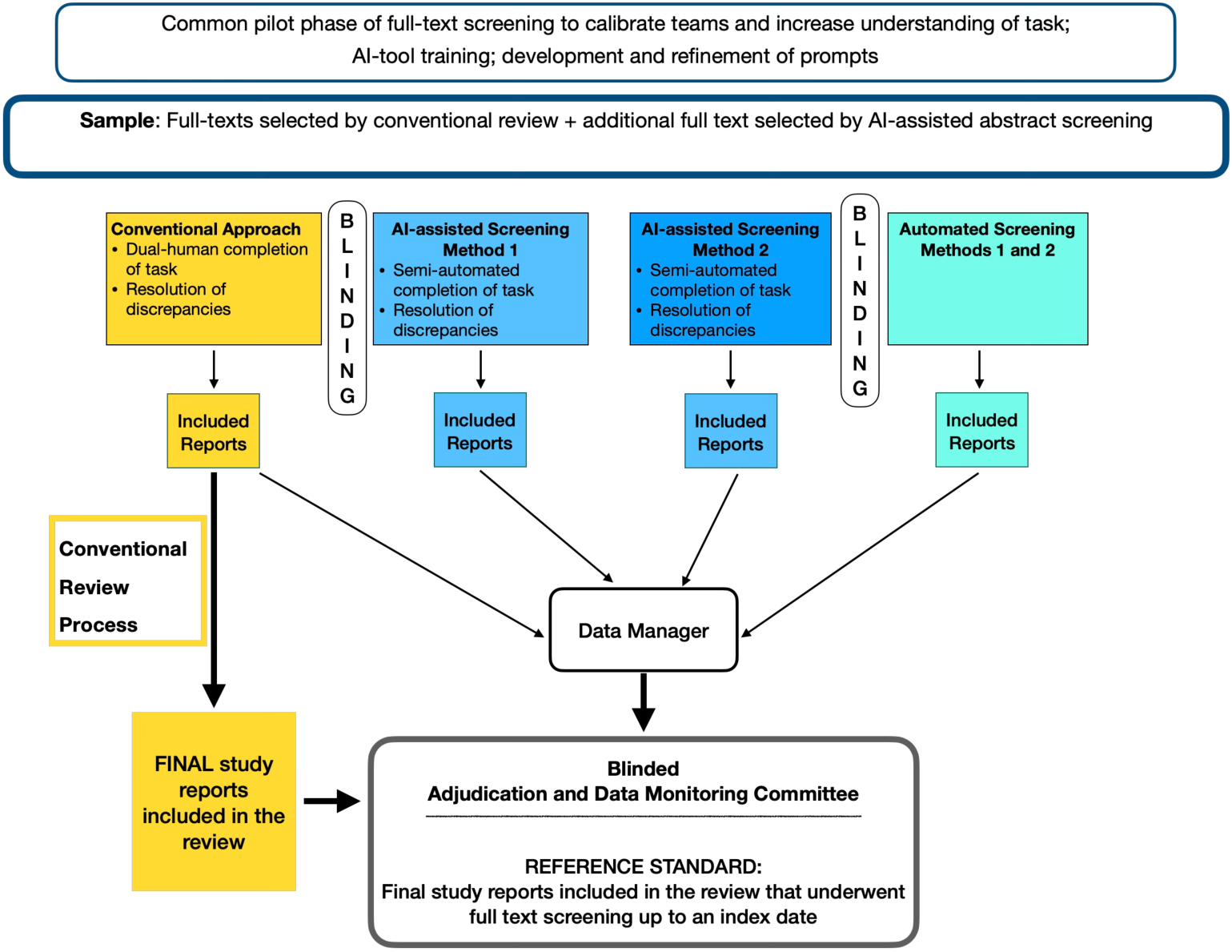
Methodological Framework for Full-text Screening Performance.

### 7.1 Sample

The sample for full-text screening will comprise all abstracts included via both the conventional approach and semi-automated abstract screening methods. We will reassess the sample definition during the interim analysis. If the workload proves excessive—for example, if the specificity of the semi-automated approaches is low—we will further restrict the sample accordingly.

### 7.2 Unit of Analysis

The unit of analysis is the study report, which is subject to full-text screening to determine whether it meets the inclusion criteria. We will keep track of studies published in multiple eligible study reports.

### 7.3 Reference Standard

The reference standard will consist of the *final included study reports* up to an index date if the update of the review has not been completed. This approach precludes human variation during full-text screening.

### 7.4 Performance Metrics

#### 7.4.1 Accuracy Metrics

We will assess the following metrics:

- *Accuracy:* The proportion of *eligible and ineligible* study reports that a full-text screening approach correctly identified
- *Sensitivity:* The proportion of the final included study reports that a full-text screening approach correctly identified
- *Specificity:* The proportion of *ineligible* study reports that a full-text screening approach correctly identified
- *Precision:* The proportion of truly eligible study reports among all reports identified as relevant
- F1 score

Exhibit B1 defines relevant accuracy metrics and components of the diagnostic 2 × 2 table for full-text screening. Proportions will be calculated with 95% Clopper–Pearson confidence intervals. For the fully automated screening approach, we will calculate the mean accuracy metrics across all iterations to assess response stability.

In addition, we will assess the following:

- *Proportion of included study reports*: The percentage of all screened records that are marked as “include” for full-text review

## Exhibit B1. Definitions of Accuracy Metrics for Full-text Screening

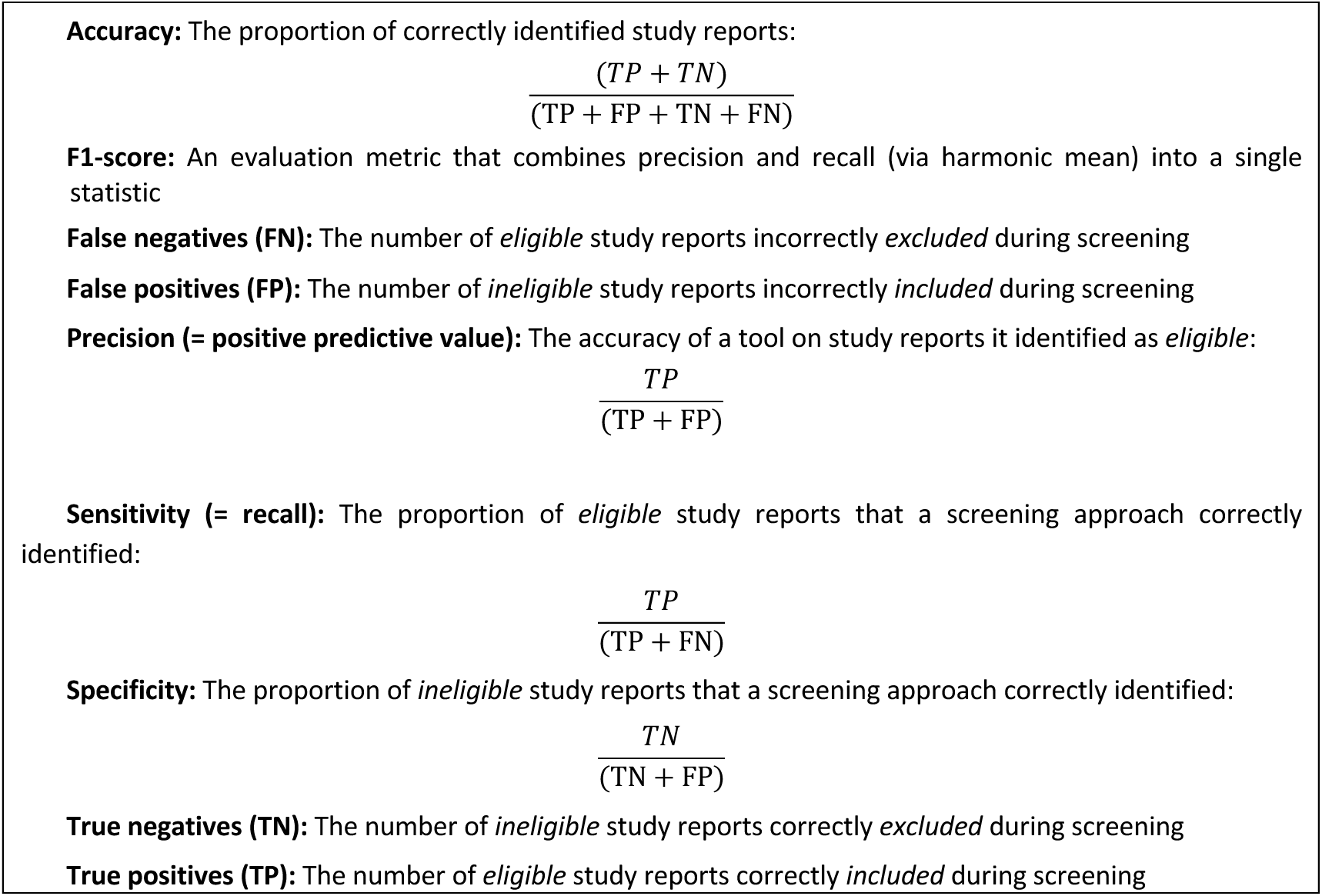

### 7.4.2 Response Stability

Refer to the master protocol.

The assessment of response stability will focus on sensitivity (recall). For each run of the tool, true positives and false negatives will be identified at the individual review level. To assess reproducibility, we will compare the sets of false negatives across runs to determine whether the same study reports are consistently falsely excluded by the fully automated approach. The overlap between false negative sets will be quantified by calculating the mean pairwise Jaccard similarity [32].

### 7.4.3 Efficiency Metrics

Refer to the master protocol.

### 7.4.4 Usability Metrics

Refer to the master protocol.

### 7.4.5 Risk Assessment and Error Analysis

Refer to the master protocol.

As with title and abstract screening, missed critical information is defined as the proportion of eligible studies incorrectly excluded during full-text screening. Changes in evidence-synthesis results will be analyzed by recalculating meta-analyses without studies incorrectly excluded at this stage.

## 8 Appendix C: Data Extraction

The evaluation of AI-assisted and fully automated data extraction will adhere to the master protocol outlined above. Figure C1 graphically illustrates the methodological framework for assessing this review task. The subsequent sections detail the methodological approaches that are specific to evaluating full-text screening.

**Figure C1.**
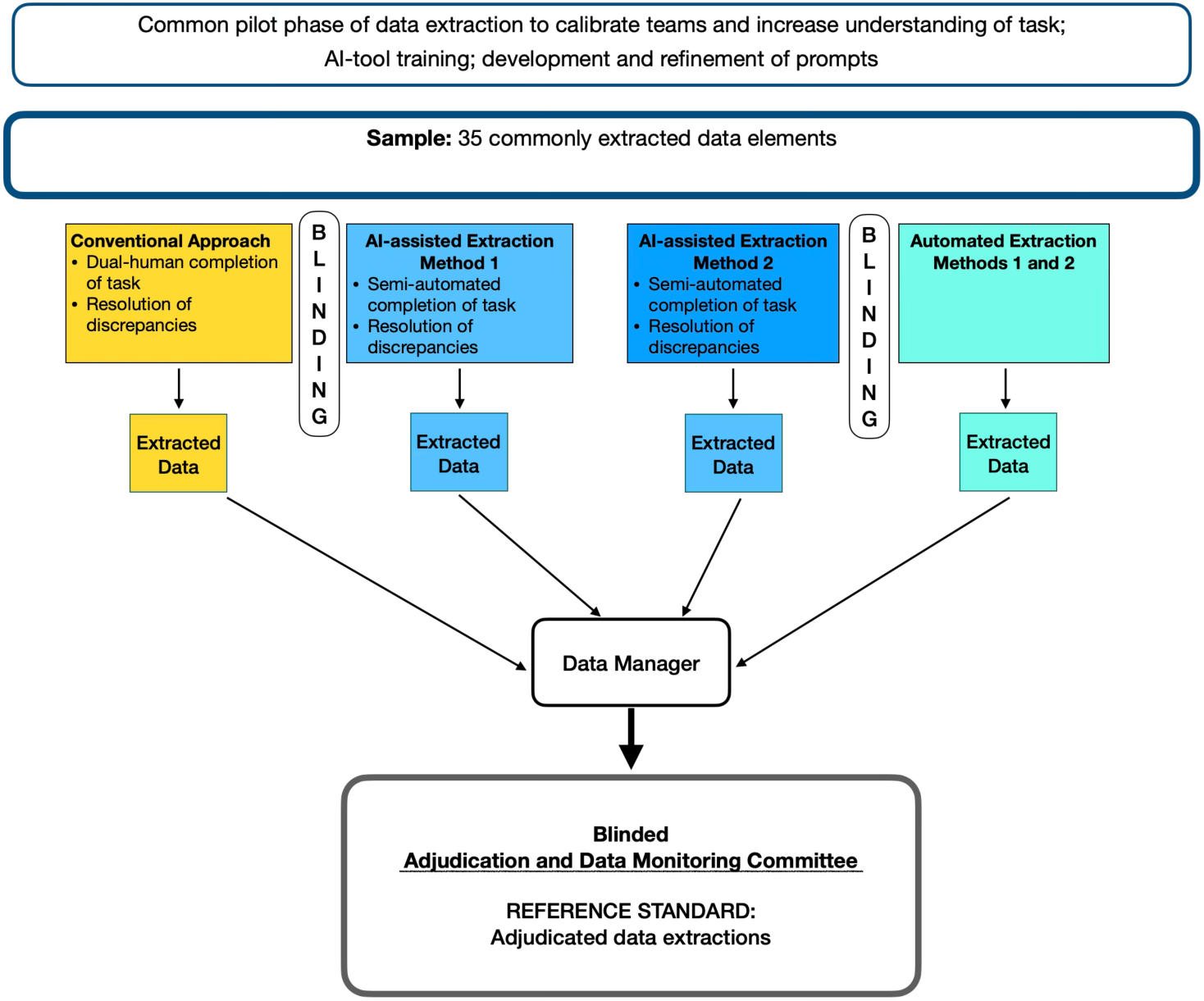
Methodological Framework for Data Extraction Performance.

### 8.1 Sample

The sample for data extraction will consist of the final included study reports from the updated Cochrane review. Review teams will provide a list of these included studies for data extraction. Data from up to ten studies, including any companion publications, will then be extracted in parallel using conventional human-only, semi-automated, and fully automated processes. If a review includes more than ten studies, we will prioritize those that report data on the primary outcomes. Should more than ten studies report on the primary outcomes, we will randomly select ten studies, regardless of the number of associated study reports. The data extraction teams will remain blinded to the data extracted by the other teams.

### 8.2 Unit of Analysis

The unit of analysis will be the individual data item (e.g., registration number, mean age of participants, count of an event). When a data item comprises multiple closely related data points—such as a point estimate and its 95% confidence interval—we will treat these as a single composite data item. Accordingly, multiple errors within a composite data item will be counted as a single error. This approach prevents artificial inflation of error counts, as these data points do not represent independent observations.

The exact unit of analysis will be determined on a case-by-case basis for each included review.

### 8.3 Selecting Study Reports for Data Extraction

Review teams will provide a list of included study reports for data extraction. Data from up to ten studies (including companion publications) will subsequently be extracted in parallel using conventional human-only, semi-automated, and fully automated processes. The data extraction teams will not be aware of the data extracted by the other teams.

### 8.4 Choice of Data Items

Due to the time constraints of this evaluation and the labor-intensive nature of adjudicating data extraction, we will focus on a sample of thirty-five extracted data items for each review. The specific data items will be determined collaboratively with the author team on a case-by-case basis when a new review is enrolled in CESAR. The data items should represent the following data categories:

- Study and publication characteristics (e.g., name of first author, study registration number, sample size)
- Participant characteristics (e.g., age, proportion of females)
- Interventions (e.g., name of interventions, dosing, route of administration)
- Participant flow (e.g., number of randomized and analyzed participants, loss to follow up)
- Statistical approach (e.g., adjustment for confounders, type of statistical analysis)
- Two primary outcomes, including one efficacy outcome and one outcome related to harms (e.g., effect estimates with 95% confidence intervals for the primary outcomes)

### 8.5 Reference Standard

Because human data extraction is error-prone, an adjudicated ground truth is required. The ADMC will resolve disagreements between data extraction approaches and determine the most accurate information for each data item. This adjudicated reference standard will then serve as the ground truth for calculating performance metrics (see below).

### 8.6 Role of the Adjudication and Data Monitoring Committee

The data manager will compile a single standardized sheet containing the outputs of all data extraction approaches, with group identifiers removed, and submit it to the ADMC. To avoid bias for or against an approach, the adjudication process will be blinded. The panel will include a senior member of the review team with topical expertise who was not involved in the data extraction task. ADMC members will first assess whether data are concordant between the two approaches. We define concordance as *data items that are factually congruent across data extraction methods, regardless of differences in style or presentation* [9]. Data items that are concordant will be assumed to reflect accurately extracted data.

Panelists will adjudicate discordant data items among approaches by consulting the original source documents to establish the “ground truth.” Data extractions will then be re-compared against the ground truth to determine whether an error is present. Finally, the panelists will judge the severity of errors in terms of their potential impact on review results and conclusions (see Table C1). A second adjudicator will review any discordances flagged by the first adjudicator and either accept the first adjudicator’s determinations or engage in dialogue to come to a consensus determination. Every final decision about the presence and severity of an error, together with the supporting rationale, will be documented to ensure a transparent and auditable trail.

**Table C1.** Types of errors in data extraction (modified from Gartlehner et al. [9])

| Type of Error | Definitions |
| --- | --- |
| Missed or omitted data | Data that were available in the study report but were either missed or omitted by the data extraction process |
| Unsupported or fabricated data | Data that were not available in the study report and were either inaccurately filled in by human data extractors or erroneously generated (hallucinated) by the AI tool |
| Misallocated data | Data that were provided in the study report but were allocated to the wrong data extraction field |
| Incorrect calculations | Mathematically incorrect calculations of data items based on information provided in the study report, including rounding errors |
| Difference in level of detail | The level of detail is the only difference |
| Ambiguous data item definition or unclear prompt | The definition of the data item lacks clarity and can be interpreted in different ways |
| Other | Optional “other” field if none of the above apply |

### 8.7 Performance Metrics

For a balanced analysis that employs a consistent denominator across all approaches, the total number of data items will remain constant. If a data item with subordinate elements is missed (e.g., if an entire outcome and its associated effect estimates or subgroup analyses are not extracted), all such items will be counted as errors, provided they are included among the thirty-five data items of interest. Although subordinate items are hierarchically dependent, we adopted this approach to prevent denominator bias.

#### 8.7.1 Accuracy Metrics

We will assess the following metrics:

- *Accuracy:* The proportion of available data items that were correctly extracted and unavailable data items that were correctly identified as such
- *Sensitivity:* The proportion of available data items that were correctly extracted
- *Precision:* The accuracy of data items for which an approach returned extracted values
- F1 score

Exhibit C1 defines relevant accuracy metrics and components of the diagnostic 2 × 2 table for data extraction. Proportions will be calculated with 95% Clopper–Pearson confidence intervals. For the fully automated screening approach, we will calculate the mean accuracy metrics across all iterations for response stability.

### Exhibit C1. Definitions of Commonly Used Terms

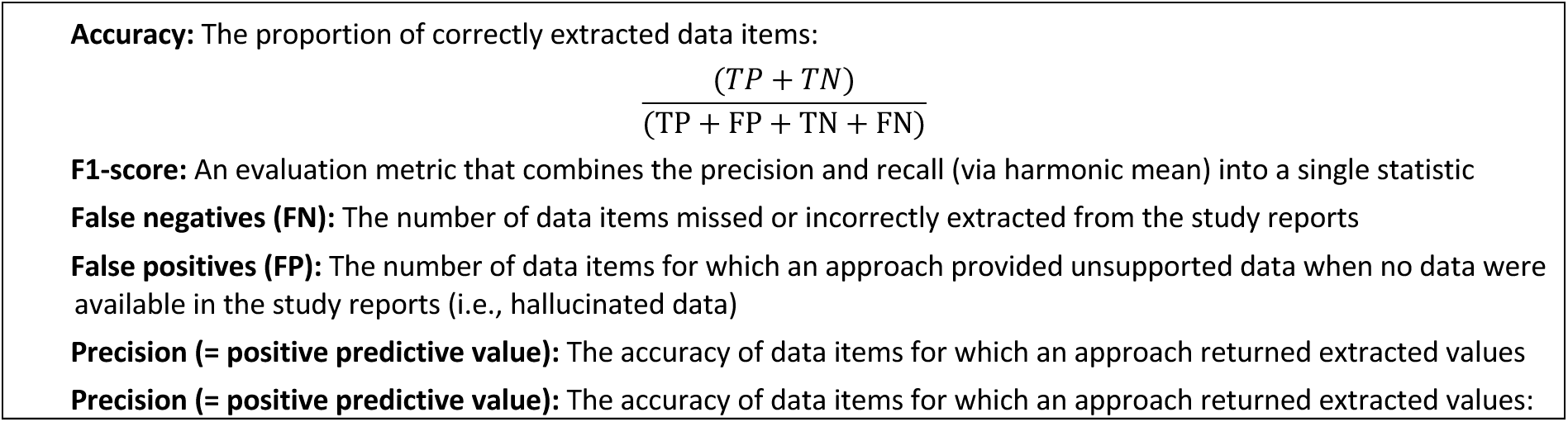
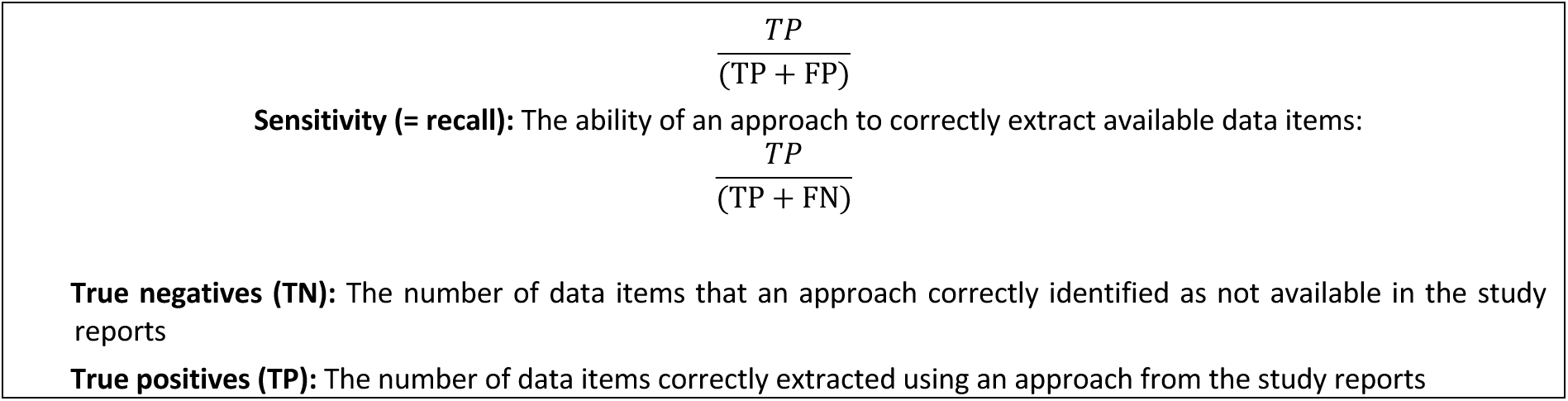

#### 8.7.2 Response Stability

Refer to the master protocol.

For data extraction, we will randomly select up to three studies per review. Automated data extraction will be performed for two numerical data items (e.g., sample size), two textual data items (e.g., names of interventions), and two mixed data items (e.g., dosing of intervention), all of which will be preselected. Due to differences in review topics, specific data items may vary between reviews. Our objective is to evaluate the overall level of agreement between runs across all data items as well as within individual data items.

#### 8.7.3 Efficiency Metrics

Refer to the master protocol.

#### 8.7.4 Usability Metrics

Refer to the master protocol.

#### 8.7.5 Risk Assessment and Error Analysis

Refer to the master protocol.

For the task of data extraction, missed critical information is defined as the prevalence of the following error types: missed or omitted data, unsupported or fabricated data, and incorrect calculations (see Table C1). Changes in evidence-synthesis results will be analyzed by recalculating meta-analyses with uncorrected errors resulting from the use of AI tools.

## 9 Acknowledgments

The authors would like to thank Sandra Hummel from Cochrane Austria for administrative support and formatting.

## 10 CRediT Authorship Contribution Statement

**G. Gartlehner:** Conceptualization, methodology, writing – original draft, writing – review & editing, project administration, supervision. **S. Banda:** Methodology, writing – original draft, writing – review & editing. **M. Callaghan:** Methodology, writing – original draft, writing – review & editing. **J. Chase:** Methodology, writing – original draft, writing – review & editing. **A. Dobrescu:** Writing – review & editing. **A Eisele-Metzger:** Writing – review & editing. **E. Flemyng:** Conceptualization, resources, writing – review & editing. **S. Gardner:** Conceptualization, methodology, writing – original draft, writing – review & editing. **U. Griebler:** Methodology, writing – original draft, writing – review & editing. **B. Helfer:** Writing – review & editing. **P. Jemiolo:** Methodology, writing – original draft, writing – review & editing. **B. Macura:** Writing – review & editing. **J.J. Meerpohl:** Conceptualization, methodology, writing – review & editing. **J Minx:** Writing – review & editing. **A. Noel-Storr:** Methodology, writing – review & editing. **N. Rajabzadeh Tahmasebi:** Methodology, writing – original draft, writing – review & editing. **A Sharifan:** Methodology, writing – original draft, writing – review & editing. **J. Thomas:** Methodology, writing – review & editing.

## 11 Funding

Part of this work was supported by the Wellcome Trust, grant number 323143/Z/24/Z, and part by the Cochrane Collaboration.

## 12 Declaration of Competing Interest

GG, who is employed at a 25% full-time equivalent by RTI International, declares an institutional conflict of interest. RTI International maintains an institutional partnership with Nested Knowledge, including an equity investment made in 2024. GG has no personal financial or non-financial ties to Nested Knowledge, receives no equity, bonuses, or remuneration from the company, and does not use Nested Knowledge in his RTI-affiliated work. GG was not involved in ranking the AI tools, which determined which AI tools entered the study.

GG, MC, AEM, EF, PJ, BM, JJM, JM, ANS, and JT are co-convenors of the joint Methods Group between Cochrane, the Campbell Collaboration, JBI, and the Collaboration for Environmental Evidence.

EF, ANS, JAC, and SG are employed by the Cochrane Collaboration.

AD and BH are engaged under a consulting agreement with Cochrane to serve as members of the Adjudication and Data Monitoring Committee.

The other authors declare that they have no conflicts of interest.

## 13 Data Availability

No data were used for the research described in this article.

## 14 Use of Generative AI

During the preparation of this manuscript, the authors utilized generative AI (Claude 4.5 Sonnet) to assist with text editing; all AI outputs were edited, where necessary, and verified by humans.

